# Ultrarapid Targeted Nanopore Sequencing for Fusion Detection of Leukemias

**DOI:** 10.1101/2022.06.20.22276664

**Authors:** Cecilia CS Yeung, Olga Sala-Torra, Shishir Reddy, Ling-Hong Hung, Jerry Radich, Ka Yee Yeung

## Abstract

Acute leukemia (AL), a cancer of the blood, has continued high mortality rates despite a plethora of available treatments. Variable prognosis, treatment response and survival are largely based on distinct cytogenetic and molecular aberrations that characterize AL subtypes. Fast and cost-effective methods of detecting AL fusions and mutations are needed to improve clinical outcomes for patients. We report preliminary data on an ultrarapid and portable nanopore long-read sequencing assay with a same-day turnaround time of ~6-7 hours for identifying key leukemic fusions. Leukemias frequently present with recurrent fusions that impact risk stratification and therapy choice, our assay can help oncologists overcome time related challenges to get patients on treatments faster.

## Introduction

Advancement of sequencing technologies have enabled large-scale and rapid molecular profiling of patients exemplified by a recent pilot study in which a genetic diagnosis of 12 critical-care patients were obtained in approximately 8-18 hours using ultrarapid whole genome sequencing^1^. Here, we report our preliminary data on a portable, ultrarapid nanopore based long-read sequencing assay with a same-day turnaround time of ~6-7 hours for identifying key leukemic fusions that can help in the management of patients. Leukemias frequently present with recurrent fusions that impact risk stratification and therapy choice^2^. Clinically defining or significant fusion genes are present in approximately 30% of acute myeloid leukemias^3^ and 30% of acute lymphoblastic leukemias^4^ These fusion genes serve a critical role driving prognosis as well as treatment, for example the use of ATRA and arsenic^5^ in acute promyelocytic leukemia and the addition of tyrosine kinase inhibitors in Philadelphia chromosome positive leukemia ^6^. To achieve our ultrarapid assay design, we combined CRISPR-cas9 based enrichment of a targeted fusion gene panel for sequencing on Nanopore flow cells with a GPU (Graphical Processing Unit)-enabled data analysis pipeline on the cloud, that allows simultaneous analyses while sequencing is occurring on the nanopore sequencer. Our integrated solution demonstrated that diagnoses can be fast and inexpensive by reducing total data generated, resources, sample requirements, and time to clinical diagnosis^7^. We present three representative cases for fusion detection in leukemia with our targeted assay and cloud-based software pipeline^8^ that takes ~3.5 hours for DNA extraction and library preparation, after which libraries are loaded directly onto the Nanopore sequencers where the portable palm sized sequencers are run attached to computers.

## Methods

Mononuclear cells were isolated using Ficoll ® reagent (Millipore-Sigma) and frozen, from primary peripheral blood or bone marrow specimens from three patients, one each with CML, APL and AML. DNA was extracted with PureGene (Qiagen, Germantown, MD, USA) following standard protocol. cRNA guides were designed to direct Cas9 to cut in the genomic proximity of each of the genes involved in each one of the translocations studied. When the target region was large, guides were tiled across the region to maximize coverage. Guides were designed to capture PML-RARA, BCR-ABL1, and KMT2A-AF4. We used 5000 ng of DNA as input and the average DNA integrity number (DIN) was 8.95 (range: 7.76-9.8). Briefly, library preparation includes an initial dephosphorylation step that renders the 5’ ends of the DNA inaccessible to adapter ligation and is followed by the addition of directional, target specific RNA guides complexed with tracrRNA and Cas9 enzyme to generate double strand DNA breaks on both ends of our region of interest. The Cas9 complex remains bound and the resulting new DNA ends contain a phosphorylated 5’end that is available for dA tailing. This results in preferential adapter ligation to these new ends. Libraries are sequenced on a MinION version 9.4. (Oxford Nanopore Technologies, Oxford, UK) sequencer where sequences are generated by DNA molecules passing through nanopores. We retrospectively took timed data packets which are binned into each read and simulated a real-time experiment where data would be streamed to our analytic pipeline on the cloud for fusion detection (where confirmation was declared when three fusions calls were seen).

## Results

Total data analysis time to confirm if a patient sample has a fusion was ~271 seconds from basecalling, alignment, fusion detection, and visualization of fusion calls. This enables a sample to fusion reporting workflow that takes an average of 5.85 hours (ranging from 5.4 to 6.3 hours), see Figure 1.

**Figure 1.**
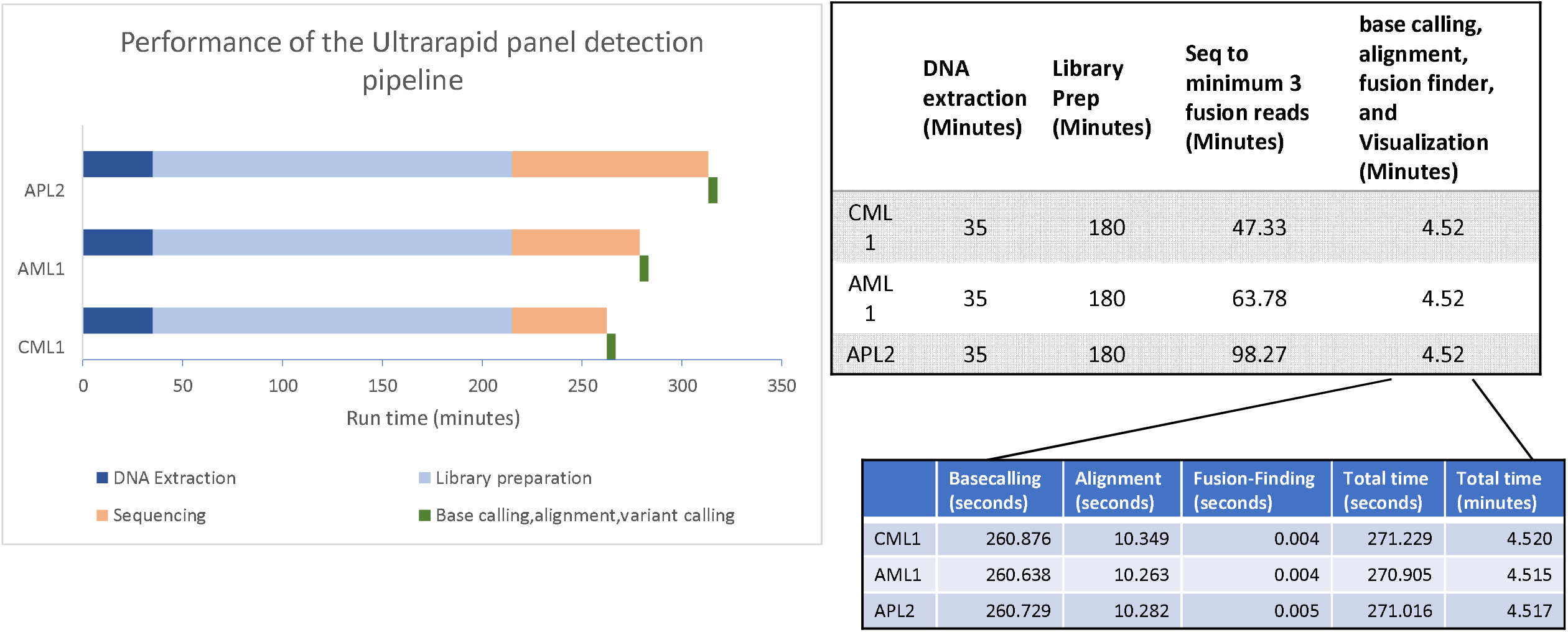
Turnaround time of our targeted, optimized assay and computational pipeline for fusion detection.

The run times in minutes of DNA extraction, library preparation, sequencing, and computational processing are shown. Reliable fusion detection is achieved when 3 fusion reads are detected. Since the basecalling, alignment and fusion detection of the first two reads occur simultaneously with sequencing, the computational times shown below the lab components include time required to basecall, align and detect the third fusion read. The basecaller’s (ONT Guppy) wrap up time is included as well. With our targeted assay, our GPU-enabled computational pipeline takes approximately 4.5 minutes after sequencing is complete. The top right grey table shows the times for sample to fusion calling charted in the bar graph (left), whereas the blue table in the bottom right corner shows a more detailed breakdown of specific times for the basecalling, alignment, fusion detection steps in our data analysis pipeline. For each of the three patients (CML, AML, APL), the total turnaround time from DNA extraction to fusion detection was 5.4 hours, 5.7 hours and 6.3 hours respectively. Our turnaround times are substantially faster than any current fusion detection assays used in clinical laboratories.

## Discussion

While our preliminary data was performed on three patients and the computational aspect of the data analysis was simulated on retrospective data, we have demonstrated the feasibility of same-day diagnosis using our ultrarapid CRISPR-case9 enrichment-based library preparation protocol paired with our cloud-based data analysis pipeline. Implemented in emergency rooms and oncology clinics, same-day diagnosis and delivery of personalized oncology treatment are now possible.

## Data Availability

Data is available for review upon request to the corresponding author and the data will be made publicly available upon acceptance.

## Declarations

The authors collectively declare that this research is conducted with approval from the Fred Hutch institutional review board and meets the requirements for ethics approval and consent to participate and consent for publication.

The authors report a provisional patent has been filed for this ultrarapid assay under (CY, OS, KYY, LHH, SR. and KY, LH has ownership in BwB as competing interests. Funding is provided in part by NCCN young investigator award and the Hyundai Hope on Wheels Scholars Award, R01 CA175008-06, UG1 CA233338-02, R01GM126019, 75N91021C00022, and 75N91020C00009.

## Authors’ contributions

CY/OS incepted the idea, OS performed all wetbench work, LHH wrote the program for BFF, SR performed informatics analysis, LH and SR created the figure, KY/CY/JR provided funding, CY drafted initial manuscript, all authors contributed to data analysis and editing of the manuscript.

## Acknowledgements

N/A

